# Association between Influenza-like illness and acute myocardial infarction patients: A case-control study

**DOI:** 10.1101/2021.04.17.21255528

**Authors:** Mehran Shokri, Seyed Morteza Mohseni, Ghasem Faghanzadeh Ganji, Arefeh Babazadeh, Kamyar Amin, Mohammad Barary, Amirhossein Hasanpour, Malihe Esmailzadeh, Soheil Ebrahimpour

**Affiliations:** Infectious Diseases and Tropical Medicine Research Center, Health Research Institute, Babol University of Medical Sciences, Babol, Iran; Student Research Committee, Babol University of Medical Sciences, Babol, Iran; Cardiac Surgery Department, Rouhani Hospital, Babol University of Medical Sciences, Babol, Iran; Department of Cardiology, Babol University of Medical Sciences, Babol, Iran; Clinical Research Development Center, Ayatollah Rohani Hospital, Babol University of Medical Sciences, Babol, Iran

**Keywords:** Influenza-like disease, Acute myocardial infarction, Vaccination, Cardiovascular diseases

## Abstract

Seasonal Influenza can cause cardiovascular complications. Therefore, this study aimed to investigate recent influenza-like illnesses (ILI) in acute myocardial infarction (AMI) patients compared to other hospitalized patients as the control group during the cold season in the north of Iran. This retrospective case-control study included 300 patients (150 AMI patients and 150 controls) aged ≥ 50 years hospitalized for acute myocardial infarction (AMI) or other conditions between September 22, 2019, and March 15, 2020. Patients in each group were frequency-matched for gender and age range. The primary exposure was a recent ILI (fever ≥ 37.8°C, cough, and sore throat) in the past month. The patients’ mean age was 64.42 ± 9.47 years, with a range of 50-94 years. Forty-five (15%) patients had diseases that met the ILI criteria. The AMI group patients significantly reported more ILI than controls (adjusted OR: 3.04, 95% CI: 1.02 to 9.09, p < 0.001). On the other hand, patients who received the influenza vaccine were significantly less likely to have an acute myocardial infarction than those who did not receive the vaccine (adjusted OR: 0.02, 95% CI: 0.001 to 0.38, p = 0.006). In conclusion, the present study demonstrates that ILI can significantly increase the risk of AMI. Also, it was confirmed that Influenza vaccination could significantly reduce the risk of AMI.

## 1. Introduction

Acute myocardial infarction (AMI) is the leading cause of death and disability [1,2]. The association between Influenza and cardiovascular events has been described and demonstrated in previous studies [3–6]. Seasonal Influenza can lead to cardiovascular complications in vulnerable individuals, especially the elderly and those suffering from underlying medical conditions [7]. First described in series of observational studies in the 1930s, evidence supports the hypothesis that seasonal Influenza may trigger AMI. Furthermore, researchers at the London Medical School have discovered that several different organisms causing respiratory infections, including the influenza virus and *Streptococcus pneumoniae*, are responsible for myocarditis which may cause blood clots in the heart muscle, increasing the chances of heart attack and stroke. New findings suggest that being infected with the mentioned organisms increases the risk of a heart attack a week after the infection and a stroke a month later [6,8–10]. Moreover, a previous study has shown that cardiovascular complications have occurred in approximately 18% of hospitalized patients due to pneumonia [11].

It is crucial to confirm the link between Influenza and AMI, as cardiovascular events induced by Influenza can be effectively prevented by vaccination. More evidence on influenza-induced cardiovascular events may lead to a change in the clinical approach that improves vaccine coverage in patients at high risk for AMI [12,13].

Considering that the association between AMI and ILI has been less studied in our country and recognizing its risk factors can pave the way for preventive interventions, it seems that a study in this regard is necessary. Therefore, this study aimed to investigate the correlation between influenza-like illnesses and acute myocardial infarction.

## 2. Material and Method

### 2.1. Study design, participants, and definition

This observational case-control study was carried out on hospitalized patients at Ayatollah Rohani Hospital in Babol, Northern Iran, from September 22, 2019, to March 15, 2020. Patients in both study groups, AMI and control, were matched for gender and age group. The age range of this study’s participants was 50-94 years. AMI group consisted of 150 patients who had experienced an AMI (defined as a rise in troponin T with ischemic symptoms, typical ECG changes, or angiographic evidence of acute coronary artery thrombosis during primary percutaneous coronary intervention). The control group encompasses 150 cases chosen from different hospital wards with no AMI history within the past month. These patients were selected as controls as their admissions were considered unlikely to be influenced by recent ILI. The primary exposure was recent ILI (fever ≥ 37.8°C, cough, and sore throat) during the past month.

### 2.2. Data collection

The recent ILI and the data on demographic characteristics, past medical history, and influenza vaccination status were collected using a data collection checklist.

### 2.3. Statistical analysis

The data were analyzed using SPSS software v.16.0 (IBM Inc., Chicago, IL, USA). Continuous and categorical variables were presented as median (IQR) and n (%), respectively. Paired t-test, Spearman’s rank-order correlation, and χ^2^ test, or Fisher’s exact test, were used to compare continuous and categorical variables. Significant values (p-values) < 0.05 were considered statistically significant.

## 3. Results

In this study, of the total 300 patients included, 150 were hospitalized in Ayatollah Rouhani Hospital’s cardiovascular ward because of the AMI, and the other 150 patients were selected from different wards of the hospital without a history of AMI. The mean age of patients was 64.42 ± 9.47 years, with a range of 50-94 years. After analyzing the demographic variables of patients admitted for AMI, a history of smoking and addiction and the presence of underlying diseases were found to be significantly associated with AMI occurrence (p < 0.001, p < 0.001, and p < 0.001, respectively). However, age, gender, and location of residence were not associated with AMI occurrence (p = 0.33, p = 0.29, and p = 0.08, respectively) (Table 1).

**Table 1:** Correlation of demographic variables in patients with and without acute myocardial infarction

| **Variables** | **Total frequency**<br>**n (%)** | **AMI group**<br>**n (%)** | **Control group**<br>**n (%)** | **p-value** |
| --- | --- | --- | --- | --- |
| **Age (years)** |  |  |  | 0.33 |
| 50-70 | 231 (77%) | 212 (74.7%) | 19 (79.3%) |  |
| ≥71 | 69 (23%) | 38 (25.3%) | 31 (20.7%) |  |
| **Gender** |  |  |  | 0.29 |
| Men | 159 (53%) | 75 (50%) | 84 (56%) |  |
| women | 141 (47%) | 75 (50%) | 66 (44%) |  |
| **Location of residence** |  |  |  | 0.08 |
| Urban | 161 (53.7%) | 73 (48.7%) | 88 (58.7%) |  |
| rural | 139 (46.3%) | 77 (51.3%) | 62 (41.3%) |  |
| **History of smoking** |  |  |  | < 0.001 |
| No | 255 (85%) | 108 (72%) | 147 (98%) |  |
| Yes | 45 (15%) | 42 (28%) | 3 (2%) |  |
| **History of addiction** |  |  |  | < 0.001 |
| No | 265 (88.3%) | 118 (78.7%) | 147 (98%) |  |
| Yes | 35 (11.7%) | 32 (21.3%) | 3 (2%) |  |
| **Underlying diseases** |  |  |  | < 0.001 |
| No | 174 (58%) | 46 (30.7%) | 128 (85.3%) |  |
| Yes | 126 (42%) | 104 (69.3%) | 22 (14.7%) |  |

Table 2 presents data on the frequency of influenza-like symptoms for the study participants. It can be inferred that muscle pain (120, 40%) was the most common symptom, followed by muscle pain (105, 35%) and chills, chills alone (33, 11%), fever and chills (18, 6%), sore throat (12, 4%), and muscle pain and diarrhea (12, 4%).

**Table 2.** Frequency of influenza-like illness symptoms in study participants

| Symptoms | Frequency<br>n (%) |
| --- | --- |
| Muscle pain | 120 (40) |
| Muscle pain and chills | 105 (35) |
| Chills | 33 (11) |
| Fever and chills | 18 (6) |
| Sore throat | 12 (4) |
| Muscle pain and diarrhea | 12 (4) |

There was also a significant association between a history of ILI and AMI occurrence (p < 0.001). Furthermore, examining the association between getting the influenza vaccine and AMI occurrence revealed a significant association between receipt of the influenza vaccine and decreased chances of AMI occurrence (p = 0.006). Moreover, patients in the AMI group were more likely to report ILI than the controls (adjusted OR: 3.04, 95% CI: 1.02-9.09, p < 0.001). The logistic regression analysis results summarized in table 3 indicated that in the analysis of unadjusted AMI variables, the age (unadjusted OR: 1.13, 95% CI: 0.75-2.23), gender (unadjusted OR: 1.27, 95% CI: 0.68-2.00), location of residence (unadjusted OR: 1.49, 95% CI: 0.94-2.19), and a history of infectious diseases (unadjusted OR: 2.04, 95% CI: 0.50-8.31) did not affect the risk of AMI occurrence.

**Table 3.** Unadjusted and adjusted odds ratios (ORs) for the correlation between acute myocardial infarction and influenza-like illnesses exposure variables

| **Exposure variable** | **AMI group**<br>**n (%)** | **Control group**<br>**n (%)** | **Unadjusted OR**<br>**(95% CI)** | **Adjusted OR**<br>**(95% CI)** | **p-value** |
| --- | --- | --- | --- | --- | --- |
| Age (50-70 years) | 212 (74.7%) | 19 (79.3%) | 1.130 (0.75-2.23) | 1.28 (0.58-2.82) | 0.33 |
| Gender (male) | 75 (50%) | 84 (56%) | 1.27 (0.68-2.00) | 1.52 (0.78-2.95) | 0.29 |
| Location of residence (urban) | 73 (48.7%) | 88 (58.7%) | 1.49 (0.94-2.19) | 1.17 (0.60-2.28) | 0.08 |
| History of smoking | 42 (28%) | 3 (2%) | 19.05 (5.75-63.09) | 15.72 (3.87-63.83) | < 0.001 |
| History of addiction | 32 (21.3%) | 3 (2%) | 13.28 (3.97-44.47) | 5.70 (1.29-25.11) | < 0.001 |
| Influenza-like illnesses | 111 (74%) | 39 (26%) | 8.43 (3.44-20.62) | 3.04 (1.02-9.09) | < 0.001 |
| Influenza vaccination | 2 (0.3%) | 148 (98.7%) | 0.15 (0.03-0.70) | 0.02 (0.001-0.38) | 0.006 |
| Infectious diseases | 6 (4%) | 3 (2%) | 2.04 (0.50-8.31) | 0.27 (0.04-1.65) | 0.31 |

This findings were also confirmed in the adjusted analysis of AMI variables’ effect, it was found that age (adjusted OR: 1.28, 95% CI: 0.58-2.82, p = 0.33), gender (adjusted OR: 1.52, 95% CI: 0.78-2.95, p = 0.29), and location of residence (adjusted OR: 1.17, 95% CI: 0.60-2.28, p = 0.08) do not affect the AMI occurrence risk. Regarding the history of infectious diseases and their effect on AMI, no significant association was observed (adjusted OR: 0.27, 95% CI: 0.04-1.65, p = 0.31).

## 4. Discussion

This retrospective case-control study aimed to investigate the correlation between Influenza-like diseases (ILI) and acute myocardial infarction (AMI) occurrence and whether Influenza vaccination can decrease AMI chances on 300 patients in Babol, Northern Iran. The most important finding of the present study was that patients with a history of ILI were at increased risk of AMI events, with the highest point estimate during the most severe influenza season.

A previous study conducted by Warren-Gash et al. in 2009 revealed that Influenza could cause AMI or cardiovascular-related mortality [9]. Several observational studies in different settings with a wide range of methods have also reported a continuing association between ILI and AMI [9,14–16].

Based on the present study’s findings, a history of smoking, addiction, and ILI were identified as risk factors for AMI which can significantly increase its chance of occurrence (p < 0.001, p < 0.001, and p < 0.001, respectively). As for smoking, its role in increasing cardiovascular events was also confirmed by other studies [17–19]. Other renowned risk factors for cardiovascular events are increased economic power of industrial societies and excessive well-being in life, excessive consumption of fats, meat, sugar, salt, and increasing tobacco consumption [20,21].

The prevalence of ILI among our patients was 50% (150 patients), of which 111 patients (74%) were in the AMI group. The most common symptoms of ILI in this study were muscle pain (120, 40%), followed by muscle pain (105, 35%) and chills, chills alone (33, 11%), fever and chills (18, 6%), sore throat (12, 4%), and muscle pain and diarrhea (12, 4%). Nevertheless, the most common ILI symptoms reported in a previous study were headaches, fever, coughing, and muscle aches [22]. Another study by Sullivan et al. demonstrated that fever, headache, sore throat, chest pain, and cough were the most prevalent ILI symptoms [23].

Furthermore, half of the patients were administered influenza vaccine before, of which 148 (98.7%) were in the control group, similar to a previous study in which 49% of patients with ischemic heart disease were vaccinated for Influenza [24]. This finding demonstrates the importance of vaccination against cardiovascular events. Another important finding of this study is that receiving the flu vaccine can significantly lower the chances of AMI, which was consistent with other studies in different parts of the world (adjusted OR: 0.02, 95% CI: 0.001 to 0.38, p = 0.006) [9,24–27].

ILI is one of the most common infectious diseases of the respiratory system, infecting many people and causing morbidity and mortality. Genetic changes in shifts and drifts in this virus are more than any other virus caused by these changes. Humans will need to produce a vaccine against the virus annually. It is necessary to find out how vaccines and antivirals are protected in patients with cardiovascular disease. Therefore, to increase influenza vaccination in patients at high risk of cardiovascular disease, a severe and intensive public health effort is necessary. The current study encompasses a short sample size. Thus, it may lack generalizability. Furthermore, subsequent clinical studies involving larger samples and more extended follow-up periods should be conducted to confirm our findings.

## 5. Conclusion

The results of the present study show that there is an association between ILI and AMI. In general, according to the findings of this study, it seems that ILI, smoking, and addiction are among the risk factors for AMI, and receiving the flu vaccine can prevent AMI occurrence.

## Data Availability

The data that support the findings of this study are available from the corresponding author upon reasonable request.

## Acknowledgments

The authors thank the Department of Infectious diseases of Babol University of Medical Sciences.

## Ethics approval statement

This study protocol was approved by the ethics committee of Babol University of Medical Sciences (IR.MUBABOL.REC.1399.031).

## Figure legend

**Figure 1.**
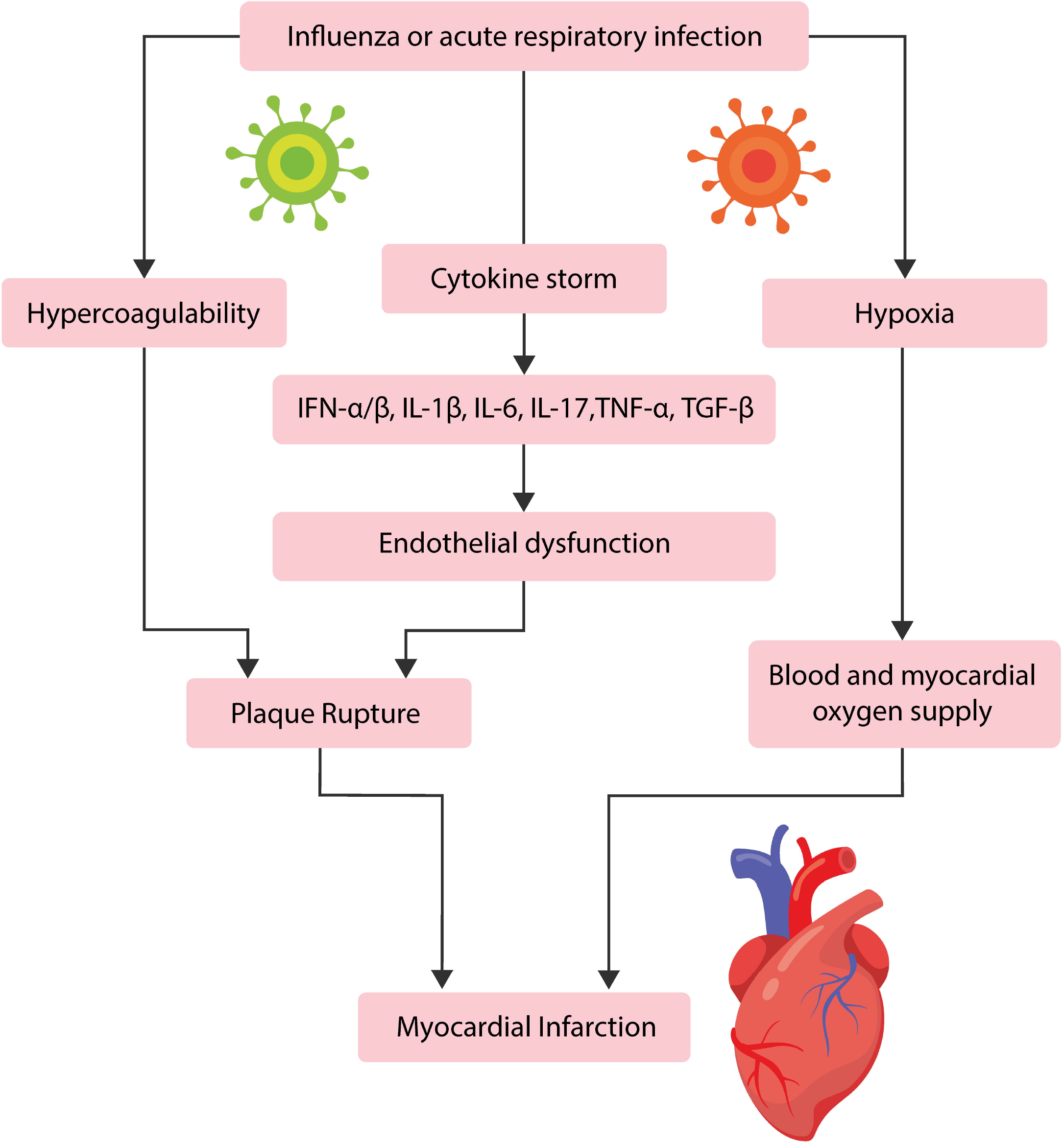
A summary of Influenza- or other acute respiratory infection-induced myocardial infarction. The Influenza virus or other acute respiratory infections can trigger three different yet highly interconnected mechanisms: Induce hypoxia, hypercoagulability state, or a cytokine storm. During a cytokine storm, interferon-alpha and -beta, interleukin-6 and -17, tumor necrosis factor-alpha, and transforming growth factor-beta can be released in high amounts causing endothelial damage. This damage, together with the hypercoagulability state, may cause plaque rupture causing a myocardial infarction. On the other hand, the induced hypoxia by those pathogens can compromise the blood and myocardial oxygen supply, which in turn, leading to myocardial infarction. Abbreviations: IFN-α, Interferon-alpha; IFN-β, Interferon-beta; IL-6, Interleukin-6; IL-17, Interleukin-17; TNF-α, Tumor necrosis factor-alpha; TGF-β, Transforming growth factor-beta

